# Serum Procalcitonin: A Novel Tumor Biomarker for Diagnosis and Follow-Up in Fibrolamellar Hepatocellular Carcinoma

**DOI:** 10.1101/2025.11.18.25340286

**Authors:** Jean-Charles Nault, Claudia Campani, Theo Z Hirsch, Ethan Neumann, Waqar Arif, Sandrine Imbeaud, Marina Baretti, Marianne Ziol, Sabrina Sidali, Pauline Roger, Manon Allaire, Mohamed Bouattour, Fabio Marra, Brice Fresneau, Neus Llarch, Jean-Marie Peron, René Gerolami, Eric Nguyen Khac, Pierre Nahon, Nathalie Ganne-Carrié, Julien Caldéraro, Aurélie Beaufrère, Valérie Paradis, Catherine Guettier, CAPRIH FFCD cohort, Angela Sutton, Mark Yarchoan, Jessica Zucman-Rossi

## Abstract

**Introduction:** Fibrolamellar carcinoma (FLC) is a rare primary liver cancer that predominantly affects young patients with normal known serum tumor biomarkers (alpha-fetoprotein (AFP) and CA19-9). An observation of a markedly elevated procalcitonin (PCT) in one patient prompted us to investigate the potential role of PCT as a biomarker in a larger cohort of FLC.

**Methods:** We measured serum PCT levels in 34 samples from 18 patients with metastatic FLC and in 64 patients with hepatocellular carcinoma (HCC), 24 with cholangiocarcinoma (CCA), and 20 with cirrhosis. Using RNA sequencing, we analyzed *CALCA* expression, the gene encoding PCT, in 27 FLC tumors, 331 HCC tumors, 39 CCA tumors, 71 hepatoblastomas, 34 hepatocellular adenomas, and 55 non-tumor liver samples. Spatial transcriptomics was performed on three FLC and PCT immunohistochemistry was conducted on 13 FLC and 34 other primary or secondary liver cancers.

**Results:** In 8 FLC from the European cohort, median serum PCT was significantly elevated (55.2 µg/l) compared to patients with HCC (0.14 µg/l), CCA (0.16 µg/l), and cirrhosis (0.11 µg/l; P=0.0005). These findings were independently validated in a U.S. cohort of 10 FLC patients compared to HCC and CCA (P=0.0002). Across these cohorts, elevated serum PCT was observed in 83% of FLC cases versus 3% of HCC and CCA cases (P<0.0001). In four patients with longitudinal measurements, changes in PCT levels correlated with radiologic response according to RECIST 1.1. RNA sequencing demonstrated significant overexpression of *CALCA* in FLC compared to other primary liver tumors (P<0.0001), and spatial transcriptomics localized *CALCA* expression specifically to tumor cells. Immunohistochemistry confirmed PCT overexpression in 77% of FLC but not in other liver cancers.

**Conclusion:** Procalcitonin is a sensitive and specific biomarker for FLC at both the serum and tumor levels among primary liver cancers, with potential utility in diagnosis and monitoring of treatment response.

**Evidence before this study:** We searched PubMed from 01th January 2000 to 03th October 2025 using the terms “fibrolamellar carcinoma”, “fibrolamellar hepatocellular carcinoma” “biomarker”, “serum”, in articles written in English Language. This analysis identified numerous studies describing the clinical, molecular, and histopathological features of fibrolamellar carcinoma (FLC), but none of them have identified a serum biomarker robustly validated for clinical use. FLC is a rare primary liver cancer typically arising in adolescents and young adults with normal liver, and current serum biomarkers used for hepatocellular carcinoma, hepatoblastoma or cholangiocarcinoma (such as alpha-fetoprotein and CA19-9) are systematically normal in FLC. Prior molecular studies have focused mainly on the *DNAJB1–PRKACA* fusion gene, which is pathognomonic for FLC, but no reliable circulating biomarker has been established for FLC diagnosis or disease monitoring.

**Added value of this study:** Our study identifies procalcitonin as a sensitive and specific biomarker for FLC, both at the serum and tumor levels. Across two independent cohorts, elevated serum PCT distinguished FLC from other primary liver cancers and from cirrhosis with high accuracy. Serum PCT level correlated with radiologic tumor response or progression, suggesting utility for disease monitoring. Transcriptomic analyses demonstrated that the *CALCA* gene, encoding PCT, is overexpressed in FLC compared with other liver tumors, and spatial transcriptomics localized *CALCA* expression specifically to tumor cells bearing the *DNAJB1–PRKACA* fusion gene. Immunohistochemistry confirmed PCT protein expression in most FLC tumors but not in other primary hepatic cancer. These findings establish a novel and readily measurable serum biomarker for FLC.

**Implications of all the available evidence:** Taken together, current evidence indicates that serum PCT is a robust diagnostic biomarker for FLC, distinguishing it from other primary liver cancers and chronic liver diseases. Routine measurement of serum PCT could facilitate earlier recognition of FLC and also provide a non-invasive tool to track treatment response. Future research should validate these findings prospectively, explore the biological mechanisms underlying *CALCA* overexpression in FLC, and assess whether PCT-guided monitoring can predict prognosis, improve patient outcomes or clinical trial design in this rare malignancy

## Introduction

Fibrolamellar hepatocellular carcinoma (FLC) is a rare primary liver cancer, with an estimated incidence of approximately 5/1000 000^1,2^. It presents with distinct clinical and biological features compared to classical hepatocellular carcinoma (HCC). While classical HCC usually arises in adult males with chronic liver disease, most commonly on cirrhosis, FLC typically develops in adolescents or young adults with normal liver and no known risk factors of chronic liver disease, and shows an equal sex distribution^3^. FLC also exhibits a unique pattern of disease spread, with frequent lymph nodes involvement, suggesting a distinct clinical behavior compared to typical HCC^4,5^. At the molecular level, FLC is defined by a characteristic chromosomal deletion of approximately 400 kilobases on chromosome 19, resulting in a chimeric fusion between *DNAJB1* and *PRKACA*^6^. This genetic alteration is considered pathognomonic for FLC and has been identified in nearly all reported cases. Functionally, the fusion protein retains the catalytic kinase domain of PRKACA thereby promoting cell proliferation^7,8^. When available in clinical practice, the identification of the *DNAJB1-PRKACA* fusion gene may aid in distinguishing FLC from other tumor subtypes with a fibrous stroma, such as HCC harboring *BAP1* mutations^9,10^.

Because of its rarity, there is currently no standard systemic therapy validated for unresectable FLC, and surgical resection remains the main treatment in cases with limited tumor burden. In contrast with classical HCC, alpha-fetoprotein (AFP) levels are normal in FLC, and no serum biomarker is available in routine clinical practice to help for diagnosis or to monitor treatment^3^.

Recently, we observed markedly elevated serum procalcitonin (PCT) levels in a patient with metastatic FLC. Procalcitonin (PCT) is coded by the *CALCA* gene and is produced at low levels by various tissues, including the liver, thyroid, lungs, and leukocytes^11,12^. It is commonly used as a biomarker for bacterial infections, as its serum concentration increases during sepsis^13^. Based on this observation, we aimed to investigate PCT expression in the serum and in tumor tissue across a larger cohort of patients with FLC as well as in other subtypes of primary liver tumors.

## Materials and Methods

### Discovery Cohort (Europe)

Patients with FLC were included in the discovery cohort based on the following criteria: histologically confirmed FLC; availability of serum and/or tumor samples; presence of active disease on imaging at the time of serum collection; and informed consent for research participation. Clinical features (age, sex, presence of extrahepatic metastasis) and biological data (serum AFP) were recorded at diagnosis (tumor sampling) and at the time of serum collection. When available, the presence of the *DNAJB1-PRKACA* fusion was recorded. The study was approved by the relevant ethics committees (HEPATOBIO CPP number CO-15-003, CAPRIH FFCD cohort CPP number 2023-A01517), and all participants provided written informed consent. The CAPRIH FFCD retro-prospective cohort is registered on ClinicalTrials.gov (NCT06541652) as well as the HEPATOBIO prospective cohort (NCT04274868).

Serum samples were collected prior to initiation of systemic therapy from 8 patients with active FLC. Control groups included 54 patients with Barcelona Clinic Liver Cancer (BCLC) stage B or C HCC and 14 patients with unresectable cholangiocarcinoma (CCA) with serums collected before the initiation of systemic treatments. In two FLC patients, sequential serum samples were collected during systemic treatment. No patients had clinical and biological signs suggestive of acute infection at the time of sampling. Serum procalcitonin (PCT) levels were measured using Abbott reagents on an Alinity analyzer (normal range < 0.5 μg/l). High-sensitivity C-reactive protein (hsCRP) was measured by an immunoturbidimetric method (reference standard ERM-DA472/IFCC) on the same analyzer (normal range < 5 mg/L).

### Validation Cohort (U.S.A)

All biospecimens included in this study were collected under oversight of the *Johns Hopkins Medicine Office of Human Subjects Research – Institutional Review Board*, located in Baltimore, Maryland, USA. The specific protocol governing the collection and use of these biospecimens is IRB00377265, titled *“Liver Cancer Biorepository to Catalyze Development of Novel Diagnostics and Treatments.”* The Johns Hopkins Medicine Institutional Review Board reviewed and approved this protocol on September 20, 2023, and the study was activated on October 19, 2023. All procedures were performed in accordance with U.S. federal regulations governing human subjects research.

The validation cohort consisted of biospecimens collected under the Johns Hopkins Biobank with the following criteria: histologically FLC with confirmation of the presence of the *DNAJB1-PRKACA* fusion; availability of serum samples; presence of active disease on imaging at the time of serum collection; and informed consent for research participation. Serum samples from FLC patients were obtained at treatment baseline from participants enrolled in a clinical trial of a therapeutic cancer vaccine for FLC (NCT04248569). Serum samples were collected from 10 patients with active FLC and measurable disease, as well as control groups of 10 HCC and 10 CCA patients. Longitudinal serum samples were analyzed in two FLC patients to evaluate dynamic changes in PCT during systemic therapy (*DNAJB1-PRKACA* fusion kinase peptide vaccine combined with Nivolumab and Ipilimumab NCT04248569). Serum PCT was measured using the Invitrogen EHPCT ELISA according to the manufacturer’s instructions. Standards were prepared in serial dilutions to generate a standard curve, and absorbance was measured at 450 nm using a microplate reader. Sample concentrations were interpolated from the standard curve, and any values below the mean absorbance of the blank control were considered undetectable and assigned a value of zero.

### Transcriptomic analysis using RNA seq

To assess the expression of the *CALCA* gene (coding for PCT), we analyzed the RNAseq data from 27 FLC, 331 HCC, 39 CCA, 71 hepatoblastoma (HB), 34 hepatocellular adenoma (HCA), and 55 non-tumor livers. Nucleic acid extraction and quality control were performed from frozen tumor and non-tumor samples as previously described^14^. mRNA sequencing (RNA-seq) was performed at Macrogen Europe and Integragen (Evry, France). We used Dragen (Illumina) for alignment and counting of the RNA-seq data. We then applied variance stabilizing transformation (VST) on the raw counts, using the Bioconductor *DESeq2* package^15^, to obtain an expression matrix without variance-mean dependence. Raw RNA-seq data (fastq format) are all publicly available in the EGA (European Genome-phenome Archive) database using the following accession numbers: EGAS00001004629;EGAS00001007694;EGAS00001007957; EGAS00001003837;EGAS00001002091;EGAS00001003685;EGAS00001005986;EGAS0000100 3310;EGAS00001005108;EGAS00001006692;EGAS00001003025;EGAS00001007348;EGAS00001002879;EGAS00001001284;EGAS00001008084 and EGAS0000100824.

### Spatial transcriptomics

Formalin-fixed paraffin embedded (FFPE) tissues from three primary FLC tumor samples were analyzed. FFPE tissue sections were cut at a thickness of 5 µm and placed on charged glass slides and were processed following the standard VisiumHD protocols from 10X Genomics (<u>CG000408</u>, <u>CG000409</u>). After Hematoxylin and Eosin (H&E) staining, the slides were assembled into Visium Cassettes and the well corresponding to the CytAssist capture area (6.5 x 6.5 mm^2^) was aligned with the area of interest. After destaining and decrosslinking, the capture area was hybridized with the Human Transcriptome Probe Set v2.0 along with a custom probe to detect the DNAJB1-PRKACA fusion transcript. The probes were released and captured onto a VisiumHD slides containing spatially barcoded probes using a CytAssist (10X Genomics). This was followed by library preparation and sequencing (∼275 million reads/sample) on an Illumina NovaSeq 6000. The sequencing reads were then aligned to the reference human genome (GRCh38) using Space Ranger v3.0 (10X Genomics) followed by merging of the 2 µm bins into segmented cells using Bin2Cell^16^. The samples were integrated with batch-effect correction and normalized using scVI (default parameters) in Python^17^. Data visualization and plot generation were performed using the Scanpy, matplotlib, and plotnine packages^18^. To assess differential expression of *CALCA* between tumor and adjacent normal cells, tumor cells were identified based on expression of the *DNAJB1-PRKACA* fusion transcript (normalized count > 0.5). Tumor and non-tumor cells were then pseudobulked, and the mean log-normalized expression of *CALCA* was computed for each group.

### Immunohistochemistry

In Europe, PCT immunostaining was performed in 8 FLC, 6 HCC with BAP1 mutations, 7 HCC without *BAP1* mutations, 7 CCA and 5 liver metastatic localization of carcinoma (colorectal, breast and serous ovarian carcinoma). Normal thyroid sample was used as a positive control. Five-micrometer-thick formalin fixed paraffin embedded were stained using an automated autostainer (Leica BondMax) with a monoclonal anti procalcitonin antibody (catalog# bsm-41284m, Bioss antibody) at a 1/200 dilution after citrate EDTA pH based epitope retrieval. Slides were scanned at X40 using an Aperio GT 450 DX Hamamatsu NanoZoomer S210.

In the USA, PCT Immunostaining was performed in 5 FLC, 4 HCC and 4 CCA at the Oncology Tissue Services and Imaging Core of Johns Hopkins University. Immunolabeling for PCT was performed on FFPE sections on a Ventana Discovery Ultra autostainer (Roche Diagnostics) using the same anti procalcitonin antibody (catalog# bsm-41284m, Bioss antibody) after epitope retrieval using CC1 Buffer. Slides were scanned at 40X using a Hamamatsu NanoZoomer S210.

Immunohistochemical slides from both cohorts were reviewed by an expert liver pathologist (MZ) and immunostaining was considered positive when 5% of more of tumor cells showed strong granular cytoplasmic staining.

### Statistical analysis

Quantitative variables were described as median (IQR= interquartile range) and dichotomous variables were described using numbers (percentages). Sensitivity, specificity and the area under the curve were used to assess the diagnostic value of serum PCT and *CALCA* mRNA level. For the comparison between quantitative variables was performed using the non-parametric Mann-Whitney test for two groups and Kruskal-Wallis or Welch’s t-test for more than two groups. Comparison of two categorical variables was performed using the Fisher exact test. The R software 4.4.3. was used to perform statistical analysis. A P value < 0.05 was considered as significant.

## Results

We incidentally identified an elevated serum PCT level in a male patient in his twenties with a recent diagnosis of FLC with peritoneal metastases. RNAseq and whole exome sequencing of the tumor confirmed the presence of *a DNAJB1-PRKACA* fusion together with a *TERT* promoter mutation. The serum PCT level was 51 μg/l (normal level < 0.5 μg/l), representing a very significant increase (102 times the upper normal limit). Such level is unusual in bacterial infections (where PCT typically ranges from 1.5 to 3 μg/l). Additionally, this patient showed no sign of bacterial infection, and the CRP level was only moderately elevated (32 ng/ml, normal < 5 ng/ml, 6-fold the upper limit of normal). Interestingly, during follow-up, the serum PCT increased to 581 μg/l (> 1000 times the normal upper limit) with confirmed radiological progression under immunotherapy.

Based on this observation, we aimed to investigate whether serum PCT could serve as a serum biomarker secreted by FLC. Initially, we measured serum PCT levels in 8 patients with FLC in Europe and compared them to levels from 14 patients with CCA, 54 with HCC, and 20 with cirrhosis without HCC (see Table 1 for the description of the patients), using a PCT assay routinely employed in France (Abbott reagents on an Alinity analyzer). The median age of FLC patients was 23 years (min=18, max=47), with 75% being male. All tumors developed on normal livers, and 75% had extrahepatic metastases. Molecular analysis was available for 4 patients, and a DNAJB1-PRKACA fusion was found in all. The median serum PCT level was 55.2 μg/l (min=0.15, max=400, IQR=26-207) in FLC, compared to 0.14 μg/l (min=0.03, max=0.65, IQR=0.09-0.28) in HCC, 0.16 μl/l (min=0.03, max=0.73, IQR=0.07-0.33) in CCA, and 0.11 μg/l (min=0.02, max=0.85, IQR=0.08-0.22) in cirrhosis (P=0.0005) (Figure 1A). Ninety percent of FLC cases (9/10) exhibited elevated serum PCT levels (above 0.5 μg/l) compared to 5% of HCC (3/54), 7% of CCA (1/14), and 5% of cirrhosis without HCC (1/20) (P<0.0001, Chi-squared test). Conversely, the median serum CRP level (normal <5 ng/ml) was 23.5 ng/ml (min=1, max=63, IQR=8-42) in FLC, versus 13 ng/ml (min=1, max=147, IQR=6-36) in HCC, 9.5 ng/ml (min=2, max=84, IQR=2-30) in CCA, and 11.6 ng/ml (min=3, max=35, IQR=9-18) in cirrhosis (P=0.7).

**Figure 1:**
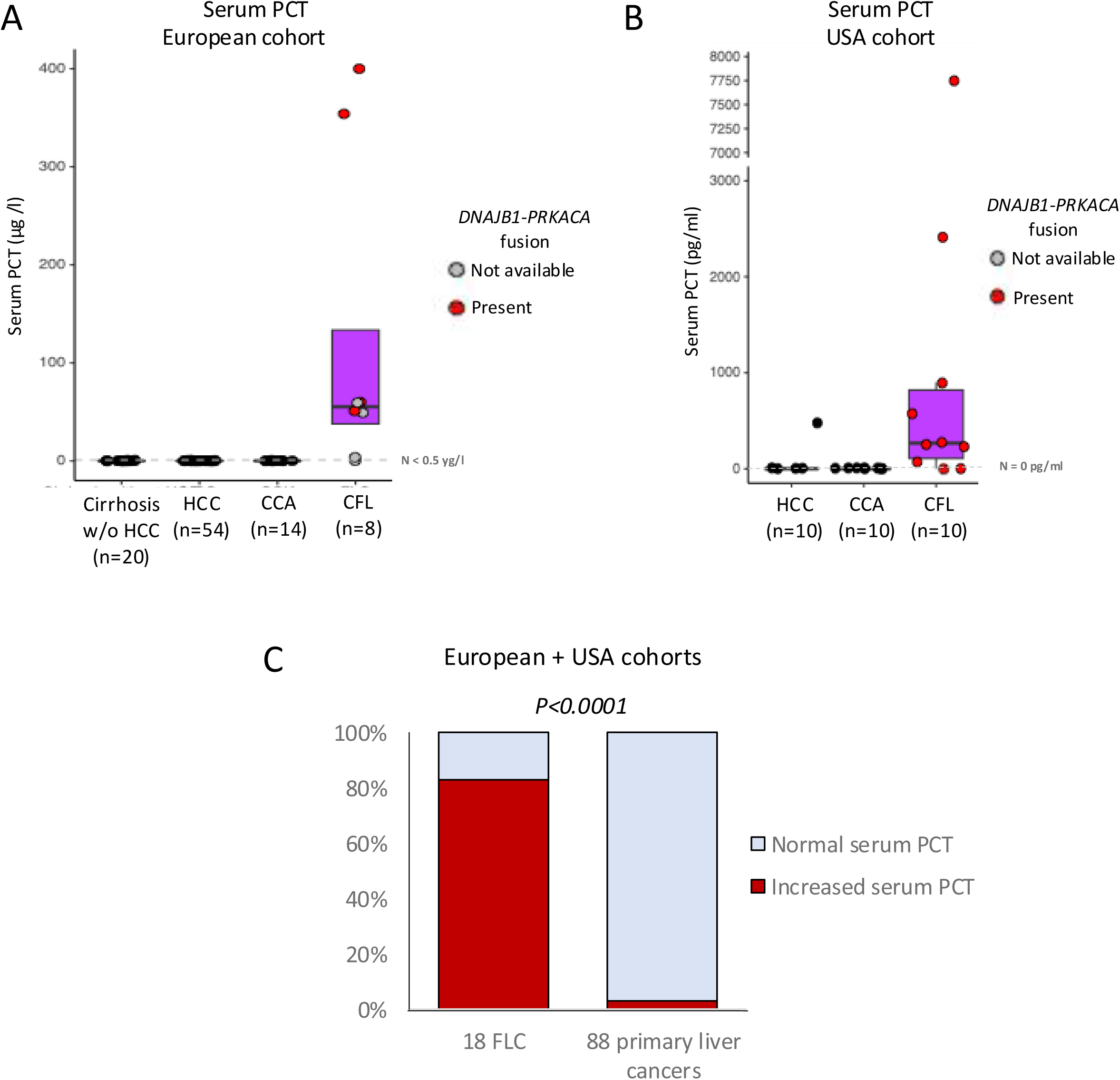
Baseline serum procalcitonin in patients with fibrolamellar hepatocellular carcinoma. A) Serum PCT levels were represented in 8 FLC compared to 14 CCA, 54 HCC and 20 cirrhosis without HCC in European patients using Abbott reagents on an Alinity analyzer. Statistical analysis was performed using the Kruskall-Wallis test. Normal level of serum PCT was below 0.5 μg/l. B) Serum PCT levels were represented in 10 FLC compared to 10 CCA and 10 HCC in patients from USA using Invitrogen EHPCT ELISA method in pg/ml. Statistical analysis was performed using the Kruskall-Wallis test. C) Distribution of positive PCT samples in 18 FLC (combination of the European cohort and USA cohort) compared to 88 primary liver cancers (24 CCA and 64 HCC). Statistical analysis was performed using Fisher exact test. Positive serum PCT was defined with a cut of of 0.5 μg/l for Abbott reagents on an Alinity analyzer (European cohort) and 0 pg/ml for Invitrogen EHPCT ELISA method (USA cohort).

**Table 1:** description of the 126 patients with available serum PCT dosage.

| ***Europe*** |  |  |  |  |
| --- | --- | --- | --- | --- |
|  | FLC<br>(n=8) | HCC<br>(n=54) | CCA<br>(n=14) | Cirrhosis without<br>HCC (n=20) |
| Age (years old) % | 23.0 (20-25) | 70.5 (63.0-78.4) | 67.6 (62.4-71.7) | 60.8 (46-70) |
| Gender (Male) <sup>§</sup> | 6 (75%) | 45 (83%) | 11 (79%) | 18 (90%) |
| Metastasis <sup>§</sup> | 6 (75%) | 14 (26%) | 8 (57%) |  |
| Non-cirrhotic liver <sup>§</sup> | 10 (100%) | 16 (29%) | 10 (71%) |  |
| Cirrhosis <sup>§</sup> | 0 (0%) | 38 (71%) | 4 (29%) | 20 (100%) |
| BCLC stage 0/A <sup>§</sup> |  | 0 (0%) |  |  |
| BCLC stage B <sup>§</sup> |  | 20 (37%) |  |  |
| BCLC stage C <sup>§</sup> |  | 34 (63%) |  |  |
| ***USA*** |  |  |  |  |
|  | FLC (n=10) | HCC (n=10) | CCA (n=10) |  |
| Age (years old) % | 25 (22-29) | 69 (73 – 65) | 57.5 (69 – 53) |  |
| Gender (Male) <sup>§</sup> | 8 (80%) | 8 (80%) | 4 (40%) |  |
| Metastasis <sup>§</sup> | 10 (100%) | 10 (100%) | 6 (60%) |  |
| Non-cirrhotic liver <sup>§</sup> | 10 (100%) | 2 (20%) | 9 (90%) |  |
| Cirrhosis <sup>§</sup> | 0 (0%) | 8 (80%) | 1 (10%) |  |
| BCLC stage 0/A <sup>§</sup> |  | 0 (0%) |  |  |
| BCLC stage B <sup>§</sup> |  | 3 (30%) |  |  |
| BCLC stage C <sup>§</sup> |  | 7 (70%) |  |  |

<sup>§</sup>Numbers (percentages) %Median (IQR)
BCLC : Barcelona liver cancer classification, FLC : fibrolamellar carcinoma, CCA: cholangiocarcinoma HCC : hepatocellular carcinoma

To validate these results, we measured serum PCT levels using a different assay (Invitrogen EHPCT) in 10 patients with FLC from the USA, and compared them with serum PCT levels in 10 patients with CCA and 10 patients with HCC (Table 1). Among the 10 patients with FLC, the median age was 22,5 years old; 80% were male; and 100% had extrahepatic metastasis. All patients had *DNAJB1-PRKACA* fusion. The median serum PCT level was 267 pg/ml (min=0, max=7444, IQR=69-901) in FLC, compared to 0 pg/ml (min=0, max=480, IQR=0-0) in HCC and 0 pg/ml (min=0, max=0, IQR=0-0) in CCA (P=0.0002) (Figure 1B). Eight of 10 FLC patients had a detectable serum PCT level (80%) compared to only one HCC patient in the control group, which included 20 primary liver cancers (5%) (P=0.0001, Fisher’s exact test).

Pooling the two cohorts (Europe and USA), an elevated serum PCT (using a cut off of > 0.5 yg/l for the European series and of > 0 pg/ml for the USA series) was detected in 83% of patients with FLC (15/18), compared with only 3% of patients with other primary liver cancers (3/88 including 64 HCC and 24 CCA) (P<0.0001, Fisher exact test) (Figure 1C). The sensitivity and specificity of increased serum PCT (using a cut off of 0.5 yg/l for the European series and a cut off of 0 pg/ml for the USA series) for the diagnosis of 18 FLC versus 87 primary liver cancers (HCC and CCA) were 83% and 96%, respectively.

We therefore analyzed the longitudinal changes in PCT levels from serum samples collected sequentially under systemic treatment in four patients with metastatic FLC (Figure 2). The first two patients were treated in Europe. The first patient showed disease progression at the initial imaging evaluation under durvalumab/tremelimumab, with serum PCT levels increasing from 51 µg/L to 207 µg/L. Progression continued under atezolizumab/bevacizumab, with PCT rising from 207 µg/L to 333 µg/L. Finally, the patient achieved stable disease with lenvatinib during which serum PCT levels fluctuated between 581 µg/L and 532 µg/L (Patient 1) (Figure 2A). The second patient initially exhibited a radiological response to lenvatinib, accompanied by a parallel decrease in serum PCT from 365 µg/L to 75 µg/L. However, secondary resistance to lenvatinib was observed, with PCT rising from 75 µg/L to 95 µg/L (Patient 2)(Figure 2B). Progression at the first imaging evaluation under second-line atezolizumab/bevazicumab was associated with a further increase in PCT from 95 µg/L to 171 µg/L.

**Figure 2:**
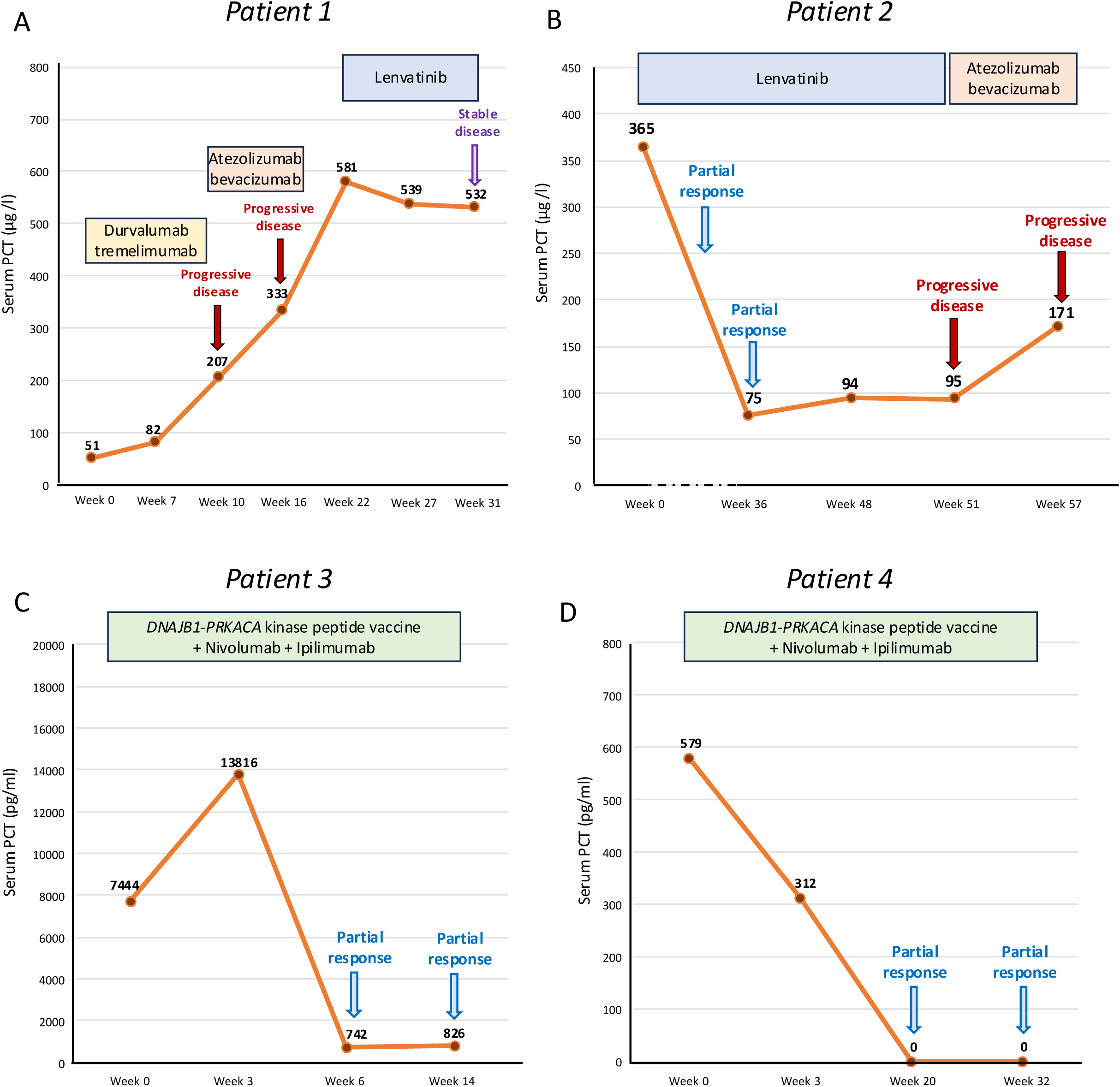
Dynamic of serum PCT levels in patients with fibrolamellar hepatocellular carcinoma under systemic treatments. We represented the evolution of serum PCT levels in four different patients with FLC receiving various types of systemic treatments (A and B in μg/l using Abbott reagents on an Alinity analyzer for the European Cohort and C and D in pg/ml using Invitrogen EHPCT ELISA method for the USA cohort). We figured the radiological response to these systemic treatments using RECIST 1.1 criteria.

The last two patients were treated in the USA by immunotherapy in a clinical trial (NCT04248569), both achieving partial radiological response. Serum PCT levels decreased from 7444 pg/ml to 826 pg/ml in one patient (patient 3) and from 570 pg/ml to 0 pg/ml (patient 4) in the other one (Figure 2C and 2D).

Next, we assessed the mRNA level of *CALCA*, the gene coding for procalcitonin, in RNAseq of 27 FLC compared to RNAseq from primary liver tumors (331 HCC without *BAP1* mutations, 21 HCC with *BAP1* mutations, 39 CCA, 71 hepatoblastoma, 14 focal nodular hyperplasia, 34 hepatocellular adenomas) and non-tumor livers (21 normal livers, 11 livers with F2-F3 fibrosis and 23 livers with F4 fibrosis) (Supplementary table 1 for the description of the samples). *CALCA* expression was significantly overexpressed in FLC compared to other primary liver tumors and non-tumor livers (P<0.0001) and, in contrast*, CALCA* was not overexpressed in *BAP1* mutated HCC, a subtype of HCC that could mimic FLC at the histological level (Figure 3A). The AUC for *CALCA* expression for the diagnosis of FLC was 1 against focal nodular hyperplasia, hepatocellular adenomas and hepatoblastomas, 0.975 against CCA and 0.994 against HCC underlying its high sensitivity and specificity for the diagnosis of this rare primary liver cancer (Figure 3B). Interestingly, *CALCA* was overexpressed specifically in tumor cells (Figure 3C) and significantly co-expressed with the *DNAJB1-PRKACA* transcript in three FLC analyzed by spatial transcriptomic, using a specific probe for fusion detection at the spatial level (Figure 3D, P=0.00276, Welch T-Test).

**Figure 3:**
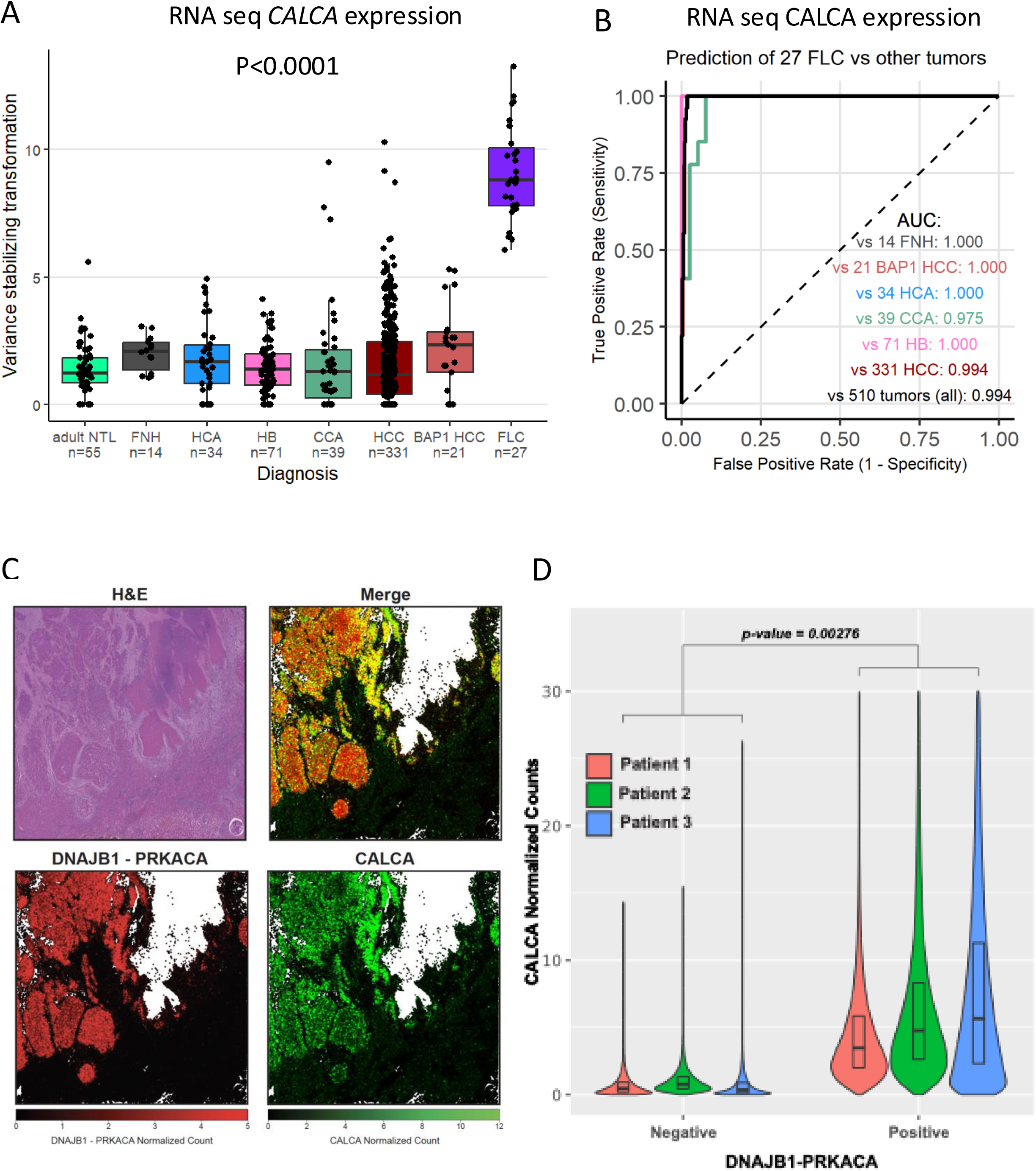
*CALCA* expression in fibrolamellar hepatocellular carcinoma. A) Expression level (Vst variant stabilizing transformation) of *CALCA*, coding for PCT, in RNA seq of 27 FLC, 21 *BAP1* mutated HCC, 331 HCC (without *BAP1* mutations), 39 CCA, 71 hepatoblastomas (HB), 34 hepatocellular adenomas (HCA), 14 focal nodular hyperplasia (FNH) and 55 non tumor livers (NTL). Statistical analysis was performed using the Kruskall Wallis test. B) Area under the curve (AUC) using *CALCA* mRNA expression for the diagnosis of FLC (n=27) versus other primary liver cancers (n=55) and non-tumor livers (21 *BAP1* mutated HCC, 331 HCC without *BAP1* mutations, 39 CCA, 71 HB, 34 HCA and 14 FNH). C) Expression of *CALCA* transcript and of the *DNAJB1-PRKACA fusion* transcrip*t* using spatial transcriptomic in one FLC. D) We represented the expression of *CALCA* between tumor and adjacent normal cells in 3 FLC. Tumor cells were identified based on expression of the *DNAJB1-PRKACA* fusion transcript. Tumor and non-tumor cells were pseudobulked, and the mean log-normalized expression of *CALCA* was computed for each group. Differential expression between tumor and non-tumor cells was assessed using a Welch’s t-test.

Finally, we assessed PCT expression by immunohistochemistry in 8 cases of FLC (7 liver resection and 1 tumor biopsy) compared with 20 cases of other primary liver tumors (6 *BAP1* mutated HCC, 7 HCC without *BAP1* mutations and 7 CCA) from Europe (Figure 4). Procalcitonin staining was also performed in 6 liver metastases of carcinoma (colorectal, breast, ovary and lung). PCT staining was positive in 7 out of 8 FLC cases and negative in all other liver cancers. In the USA, PCT expression by immunohistochemistry was assessed in 5 cases of FLC and 8 primary liver cancers (4 HCC, 4 CCA) and PCT was positive in 3/5 FLC but in none of the 8 primary liver cancers. Pooling the data from Europe and USA, PCT staining was positive in tumor cells in 10 FLC cases among 13 (77%) versus none (0%) of the 28 primary liver cancers (P<0.0001, Fisher-exact test) (Figure 4A and 4B). As shown in figure 4A and 4B, the tumors showed an heterogenous pattern consisting of several groups of tumor cells with a strong granular cytoplasmic staining. The percentage of positive tumor cells ranged from 5 to 50% in FLC (Figure 4C, 4D and AE). Non-tumoral liver did not show any significant staining.

**Figure 4:**
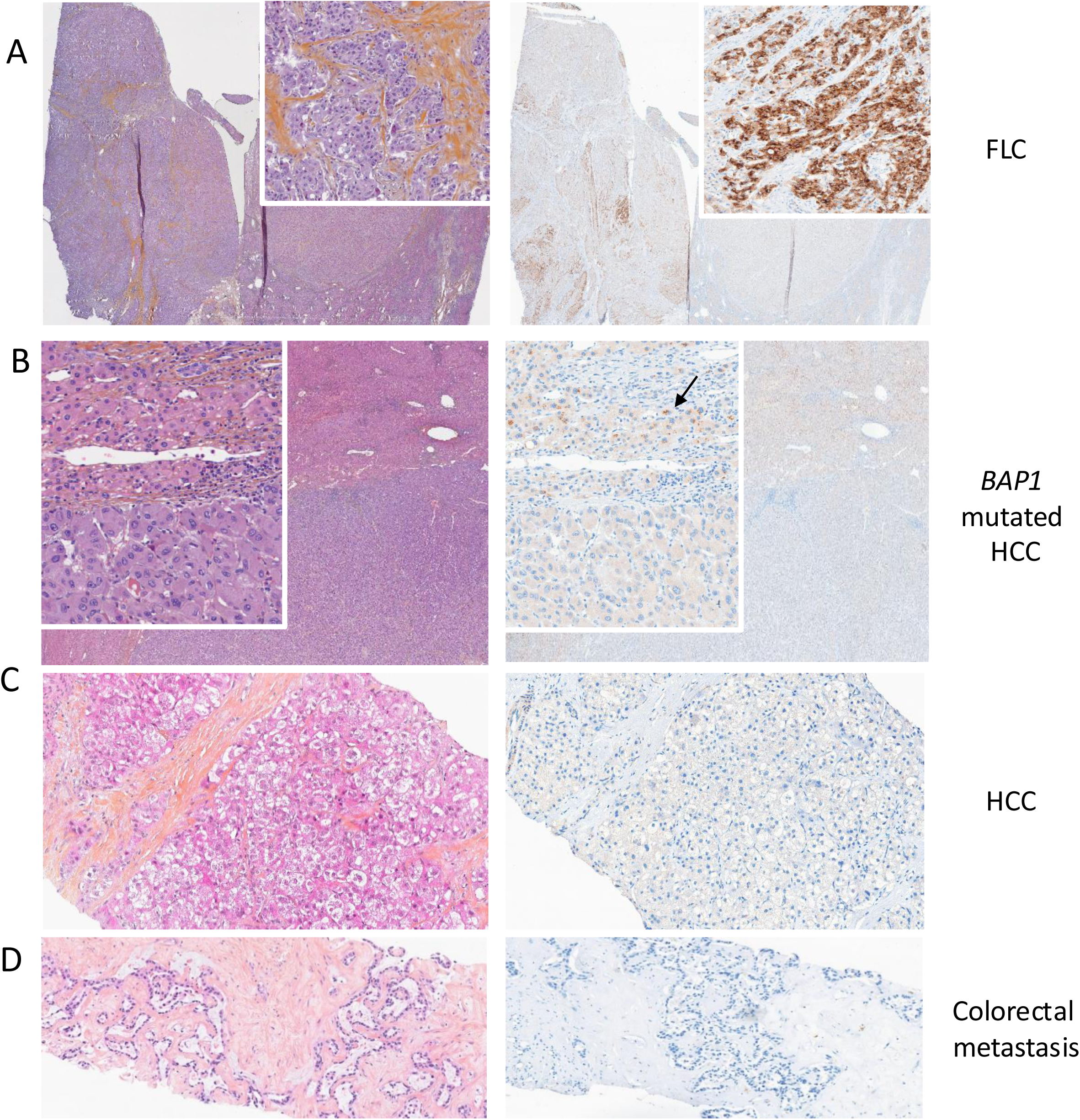
PCT immunohistochemistry in fibrolamellar hepatocellular carcinoma and other primary liver cancers. PCT immunohistochemistry in (A) fibrolamellar carcinoma, (B) *BAP1* mutated HCC, (C) HCC without *BAP1* mutations and (D) CCA. Fibrolamellar carcinoma resection sample showed several areas of tumor cells with strong granular cytoplasmic staining. The staining was heterogeneously distributed with positive and negative areas (line A, lox and high-power field inset, hematein eosin and saffron left, procalcitonin staining right). Hepatocellular carcinoma with *BAP1* mutation resection sample did not show any positive tumor cell (line B, low and high-power field inset, hematein eosin and saffron left, procalcitonin staining right). Brown staining in non-tumor hepatocyte (arrow) corresponds to lipofusin (line B, low and high-power field inset, hematein eosin and saffron left, procalcitonin staining right). No hepatocellular carcinoma (line C) and cholangiocarcinoma (line D) biopsy samples did not show any positive tumor cells.

## Discussion

The identification of specific and sensitive serum tumor biomarkers remains a critical area of research in the field of rare primary liver tumors since AFP and carbohydrate CA 19-9 are currently the only serum tumor biomarkers routinely used for primary liver tumors, specifically for HCC/Hepatoblastomas and CCA, respectively^19,20^. In our study, we identified PCT as a serum tumor biomarker for metastatic FLC increased in 80% of these patients. Serum PCT also proved useful for monitoring patients undergoing treatment for FLC, serving as a tumor-secreted biomarker.

The *CALCA* gene encodes procalcitonin, which is primarily cleaved into calcitonin in thyroid C-cells, where it regulates calcium homeostasis^12^. Consequently, medullary thyroid carcinoma is characterized by paraneoplastic secretion of calcitonin, which is widely used for diagnosis and treatment monitoring^21^. In other tissues, *CALCA* gene expression is minimal and typically upregulated in response to bacterial infections. However, outside the thyroid, procalcitonin cannot be processed into calcitonin. As a result, serum PCT levels are commonly used as a biomarker for bacterial infections, with diagnostic thresholds typically exceeding 0.5 µg/L, but exceptionally rising above 20 µg/L^22,23^. However, in our study, the level of serum PCT identified reaches a very high level in patients with our series and appears to be both highly sensitive in FLC, as it is virtually absent in other primary liver tumors, whether benign or malignant. Interestingly, in our study, serum PCT was overexpressed in 83% of patients with FLC. These results were identified in a European cohort of patients with FLC and externally validated in a USA cohort of FLC patients. In the literature, only one case of increased PCT has been previously reported in one patient with FLC^24^. Importantly, the usage of 2 different methods of quantification of PCT in Europe and in the USA showed convergence in elevated PCT despite a different range of values. This highlights the need to define diagnostic thresholds specific to the dosage method in future studies.

We acknowledge that the number of available serum samples was limited due to the extremely rare incidence of FLC. To further support the specificity of this biomarker, we analyzed *CALCA* mRNA expression in a large and diverse cohort of primary liver tumors as well as non-tumoral liver tissues (normal, fibrotic, and cirrhotic liver). We found that only FLC exhibited overexpression of *CALCA* mRNA at RNA seq with an AUC from 0.994 to 1. Overexpression of *CALCA* has previously been reported in FLC, alongside other genes associated with neuroendocrine phenotype, such as *PCSK1*, *NTS*, and *DNER*^25,26^. Increased serum PCT have been previously identified in cancers such as medullary thyroid carcinoma or, more rarely, in some cases of small-cell lung cancer, underlying the link between neuroendocrine features and PCT overexpression^25,26^ However, the specificity of *CALCA* overexpression in FLC, and the potential to detect its protein product PCT in patients’ serum and tumors for treatment monitoring, had not been demonstrated before our study.

We also validated PCT expression at the protein level using immunohistochemistry, which demonstrated overexpression in FLC but not in other primary liver tumors. These findings suggest that PCT immunostaining could serve as a useful diagnostic tool to confirm FLC histologically, especially in settings where molecular detection of the *DNAJB1–PRKACA* fusion is not readily available in clinical practice^9^. Moreover, a potential diagnostic challenge arises with *BAP1*-mutated HCC, which may exhibit fibrolamellar features and often develops in non-cirrhotic livers, similar to FLC^14,27^. However, these tumors typically occur in older patients and present with distinct clinical behavior, including fewer lymph node metastases. In our study, PCT was not overexpressed at the mRNA or protein level in *BAP1*-mutated HCC, making it a potentially valuable biomarker for distinguishing these tumors from FLC.

One potential limitation is that serum PCT can also be elevated in the context of bacterial infections^12^. However, active bacterial infection can typically be ruled out based on routine clinical and biological assessment. Furthermore, the median serum PCT levels in patients with bacterial infections are significantly lower than those observed in patients with FLC. In randomized controlled trials of septic patients, median PCT levels were reported as 1.6 µg/L (IQR: 0.5–6.6) and 1.9 µg/L (IQR: 0.40–14.1), whereas PCT levels in patients with FLC often exceed 10 µg/L^22,23^. In these trials, the method of PCT dosage was similar to what we used in the European cohort of FLC. Additionally, serum C-reactive protein (CRP) lacks sensitivity as overexpression is not constant in patients with FLC but also of specificity, as similar elevations are also observed in advanced HCC, metastatic CCA, and inflammatory hepatocellular adenomas^28^. Finally, most of patients of our study had metastases or locally advanced FLC and serum PCT levels should be assessed in patients with lower tumor burden accessible to liver resection.

In conclusion, we identified a novel serum tumor biomarker, procalcitonin, that is highly specific for FLC among all primary liver cancers and increased in 80% of patients with metastatic FLC. This biomarker holds promise for improving the diagnosis of this rare tumor and for monitoring treatment response.

## Data Availability

All RNA seq data are publicly available in EGA (see materials and methods, EGA number: EGAS00001008247)

## Aknowledgment

We are deeply grateful to the patients and their families for their contribution of biospecimens to this work. We would like to thank all the investigators of the CAPRIH FFCD cohort coordinated by the FFCD (Federation Française de Cancérologie Digestive), Lila Gaba, Daniel Gonzalez and Karine Le Malicot and the Johns Hopkins Liver Cancer Biobank who donated biospecimens for cancer research, all the investigators of the HEPATOBIO consortium, Angèle Priso as well as all the members of the Paris Liver Cancer Group (PLCG), and the healthcare providers who cared for the participant via the Johns Hopkins Multidisciplinary Liver Cancer Clinic.

---

**Supplemental table 1:** description of the 592 patients with available tumor or non-tumor RNA seq analyzed.

|  | **FLC**<br>**(n=27)** | **HCC**<br>**(n=331)\*** | **BAP1 HCC**<br>**(n=21)** | **CCA**<br>**(n=39)** | **HB**<br>**(n=71)** | **FNH**<br>**(n=14)** | **HCA**<br>**(n=34)** | **NTL**<br>**(n=55)** |
| --- | --- | --- | --- | --- | --- | --- | --- | --- |
| **Age%** | 19<br>(14-27) | 62<br>(52-70) | 51<br>(41-61) | 65<br>(55-75) | 2.0<br>(1.0-5.0) | 33<br>(28, 42) | 32<br>(25-45) | 57<br>(44-70) |
| **Male** | 12/27<br>(44%) | 273/331<br>(82%) | 6/21<br>(29%) | 24/38<br>(63%) | 46<br>(65%) | 4/14<br>(29%) | 29/34<br>(85%) | 38/55<br>(69%) |
| **Chronic Alcohol intake<sup>§</sup>** | 0/27<br>(0%) | 67/326<br>(21%) | 4 /18<br>(22%) | 3/38<br>(8%) | 0<br>(0%) |  | 2/34<br>(6%) | 6/55<br>(11%) |
| **Chronic Hepatitis B<sup>§</sup>** | 0/27<br>(0%) | 129/326<br>(40%) | 2/18<br>(11%) | 1/38<br>(3%) | 0<br>(0%) |  | 0<br>(0%) | 25/55<br>(45%) |
| **Chronic Hepatitis C<sup>§</sup>** | 0/27<br>(0%) | 44/326<br>(13%) | 0/18<br>(0%) | 4/38<br>(11%) | 0<br>(0%) |  | 1/34<br>(3%) | 7/55<br>(13%) |
| **Metabolic syndrome<sup>§</sup>** | 1/27<br>(5.3%) | 10/326<br>(3.1%) | 1/18<br>(5.6%) | 2/38<br>(5%) | 0<br>(0%) |  | 4 (25%) | 4/55<br>(7.3%) |
| **Fibrosis NTL F0-F1<sup>§</sup>** | 27/27<br>(100%) | 109/329<br>(33%) | 17/18<br>(94%) | 11/34<br>(32%) | 63/71<br>(100%) | 12/14<br>(86%) | 25/29<br>(86%) | 21<br>(38%) |
| **Fibrosis NTL F2-F3<sup>§</sup>** | 0/27<br>(0%) | 81/329<br>(25%) | 0/18<br>(0%) | 21/34<br>(62%) | 0<br>(0%) | 1/14<br>(7%) | 3/29<br>(10%) | 11<br>(20%) |
| **Fibrosis NTL F4<sup>§</sup>** | 0/27<br>(0%) | 139/329<br>(42%) | 1/18<br>(6%) | 2/34<br>(5.9%) | 0<br>(0%) | 1<br>(7%) | 1/29<br>(3.4%) | 23<br>(42%) |
| **Serum AFP level<sup>%</sup>** | 1.1<br>(1-1) | 12 (4, 55) | 59<br>(16-145) | 1.1<br>(1-1) | 42<br>(17, 130) |  |  |  |
| **BCLC stage 0<sup>§</sup>** |  | 12<br>(4.0%) | 8/14<br>(57%) |  |  |  |  |  |
| **BCLC stage A<sup>§</sup>** |  | 183<br>(60%) | 4/14<br>(29%) |  |  |  |  |  |
| **BCLC stage B<sup>§</sup>** |  | 61<br>(20%) | 0/14<br>(0%) |  |  |  |  |  |
| **BCLC stage C<sup>§</sup>** |  | 47<br>(16%) | 2<br>(14%) |  |  |  |  |  |

\*HCC without *BAP1* mutations <sup>§</sup>Numbers (percentages) <sup>%</sup>Median (IQR)
BCLC : Barcelona liver cancer classification, FLC : fibrolamellar carcinoma, HB : hepatoblastoma, HCC : hepatocellular carcinoma, FNH : focal nodular hyperplasia, NTL: non-tumor liver

